# Diagnostic Validity of Drinking Behaviour for Identifying Alcohol Use Disorder: Findings from a Nationally Representative Sample of Community Adults and an Inpatient Clinical Sample

**DOI:** 10.1101/2024.09.14.24313683

**Authors:** M.L. Garber, A. Samokhvalvov, Y. Chorny, O. LaBelle, B. Rush, J. Costello, J. MacKillop

## Abstract

**Background and Aims:** Alcohol consumption is an inherent feature of alcohol use disorder (AUD), and drinking characteristics may be diagnostically informative. This study had three aims: (1) to examine the classification accuracy of several drinking quantity/frequency indicators in a large representative sample of U.S. community adults; (2) to extend the findings to a clinical sample of adults; and (3) to examine potential sex differences.

**Design:** This retrospective study utilized receiver operating characteristic (ROC) curves to evaluate area under the curve (AUC). Optimal cut-offs were identified using the Youden Index. Diagnostic validity was evaluated using accuracy, sensitivity, specificity, positive predictive value (PPV), and negative predictive value (NPV).

**Measurements:** Index tests included measures of quantity/frequency (e.g., drinks/drinking day, largest drinks/drinking day, number of drinking days, and heavy drinking frequency). The reference standard was AUD status as determined via a clinical interview (community sample) or a symptom checklist (clinical sample).

**Setting and Participants:** Two samples were examined: A large, nationally representative random sample of U.S. community adults who reported past-year drinking (*N*=25,778, AUD=20%) and a consecutive drinking clinical sample from a Canadian mental health and addictions inpatient treatment centre (*N*=1,341, AUD=82%).

**Findings:** All drinking indicators performed much better than chance at classifying AUD (AUCs=0.60-0.92, *p*s<.0001). Heavy drinking frequency indicators performed optimally in both the community (AUCs=0.78-0.87; accuracy=0.72-0.80) and clinical (AUC=0.85-0.92; accuracy =0.77-0.89) samples. Collectively, the most discriminating drinking behaviors were heavy drinking episodes and exceeding NIAAA drinking low-risk guidelines. No substantive sex differences in optimal cut-offs or variable performance were observed.

**Conclusions:** Drinking patterns performed well at classifying AUD in both a nationally representative and large inpatient sample, robustly identifying AUD at rates much better than chance and above accepted benchmarks, with limited differences by sex. Findings broadly support the potential utility of quantitative drinking indicators as being diagnostically informative in clinical settings.

## INTRODUCTION

Diagnosis of psychiatric conditions are complicated by the absence of a singular objective biological marker. Instead, diagnosis in psychiatry typically relies on a constellation of behavioural and cognitive symptoms outlined by two major classification systems: the International Classification of Diseases (ICD) and Diagnostic and Statistical Manual of Mental Disorders (DSM-5-TR) (1,2). Conditions are identified via clinical interviews conducted by healthcare professionals, which are the gold standard in psychiatric diagnosis. Alcohol use disorder (AUD), and substance use disorders more broadly, are somewhat unique in that an overt, measurable behaviour (i.e., substance use) is necessary for a diagnosis to be made. That is, AUD cannot be present in the absence of drinking behavior. Despite this, somewhat paradoxically, diagnostic definitions do not include patterns of alcohol consumption. Indeed, some researchers have argued that many SUD diagnostic criteria are necessarily defined by heavy substance use over time, sparking debate about whether “heavy use over time,” or similar behavioral indicators are sufficient for identifying AUD (3–9).

A modest number of empirical studies have explored the relationship between drinking quantity and frequency variables and AUD criteria. For example, one study examined the effect of including a drinking pattern criterion (drinking more than 5 drinks per occasion for males, more than 4 drinks per occasion for females) along with other DSM diagnostic criteria. Using item response theory analyses, they found that the drinking pattern criterion was related to AUD status and severity, particularly lower severity AUD (10). Another investigation found that in a treatment seeking sample of young adults, consumption variables contributed to measurement information beyond AUD criteria across the severity spectrum, and that combining AUD symptoms with indices of alcohol consumption better predicted alcohol involvement after treatment than AUD symptom counts or a DSM-IV dependence diagnosis alone (11). These studies highlight relationships between drinking consumption and AUD status, as well as the potential clinical utility of consumption measurement. However, they do not address whether consumption measures can accurately identify AUD. An assessment tool that utilizes consumption characteristics with a validated cut-off may serve useful for several reasons. First, consumption variables are brief to administer and can be easily integrated into standard clinical practice with minimal clinician and patient burden. Second, alcohol consumption is an easily collected, objective, and quantifiable behaviour. This may help determine whether a more time-intensive, detailed, and rigorous assessment for an AUD diagnosis is warranted.

Some studies have examined the utility of drinking quantity/frequency as a screening tool for classifying AUD status. One recent study examined the accuracy of a frequency measure (number of drinks per week over the previous year) in classifying AUD status in a sample of young Swiss men (12). They found that a cut-off of 21 or more drinks per week achieved a 75% accuracy rate, but that sensitivity was extremely poor (0.39). On the other hand, the highest sensitivity and specificity (0.83 and 0.60, respectively) was achieved with a cut-off of 10.5 drinks per week or more, but the accuracy rate associated with this cut-off was only 65%.

Furthermore, several studies have examined the screening properties of the *Alcohol Use Disorder Identification Test – Consumption* (AUDIT-C), a composite of three drinking quantity/frequency items used as a screening tool for AUD (13). These studies have generally found that the AUDIT-C is an accurate screener for AUD status, with acceptable to excellent sensitivity and specificity (14–17). However, the AUDIT-C does not provide a cut-off for a specific level of drinking quantity or frequency. Rather, it is a composite of three binned quantity and frequency items and does not inform the accuracy of a singular measure of behaviour at classifying AUD status.

The literature examining relationships between alcohol consumption and AUD status suggests the potential promise of using drinking quantity/frequency indicators as an assessment tool for classifying AUD. However, few studies have specifically examined the classification accuracy of quantity and frequency variables, and existing studies have primarily examined the AUDIT-C, a composite of several quantity/frequency measures. The current study sought to extend the existing literature by examining the classification accuracy of alcohol consumption characteristics in two samples: (1) a large, nationally representative sample of U.S. community adults, and (2) a sample of adults seeking treatment for substance use disorders in an inpatient treatment setting in Ontario, Canada. Additionally, this study evaluated possible sex differences.

## METHOD

### Participants

#### Community Sample

This study utilized data from the National Epidemiological Study on Alcohol and Related Conditions Study III (NESARC-III). NESARC-III comprises a nationally representative and randomly selected sample of community adults from the United States. Participants were included in the current study’s analyses if they reported drinking over the past 12 months (*N* = 25,778; 71% of total sample), had been assessed for AUD, and responded to questions about the frequency/quantity of their past year alcohol intake. Among those with AUD, most had a mild condition (18). See Table 1 for participant characteristics and Figure 1 for the STARD diagram. For more detailed information about data collection methods, see Grant et al., 2014 (19).

**Figure 1:**
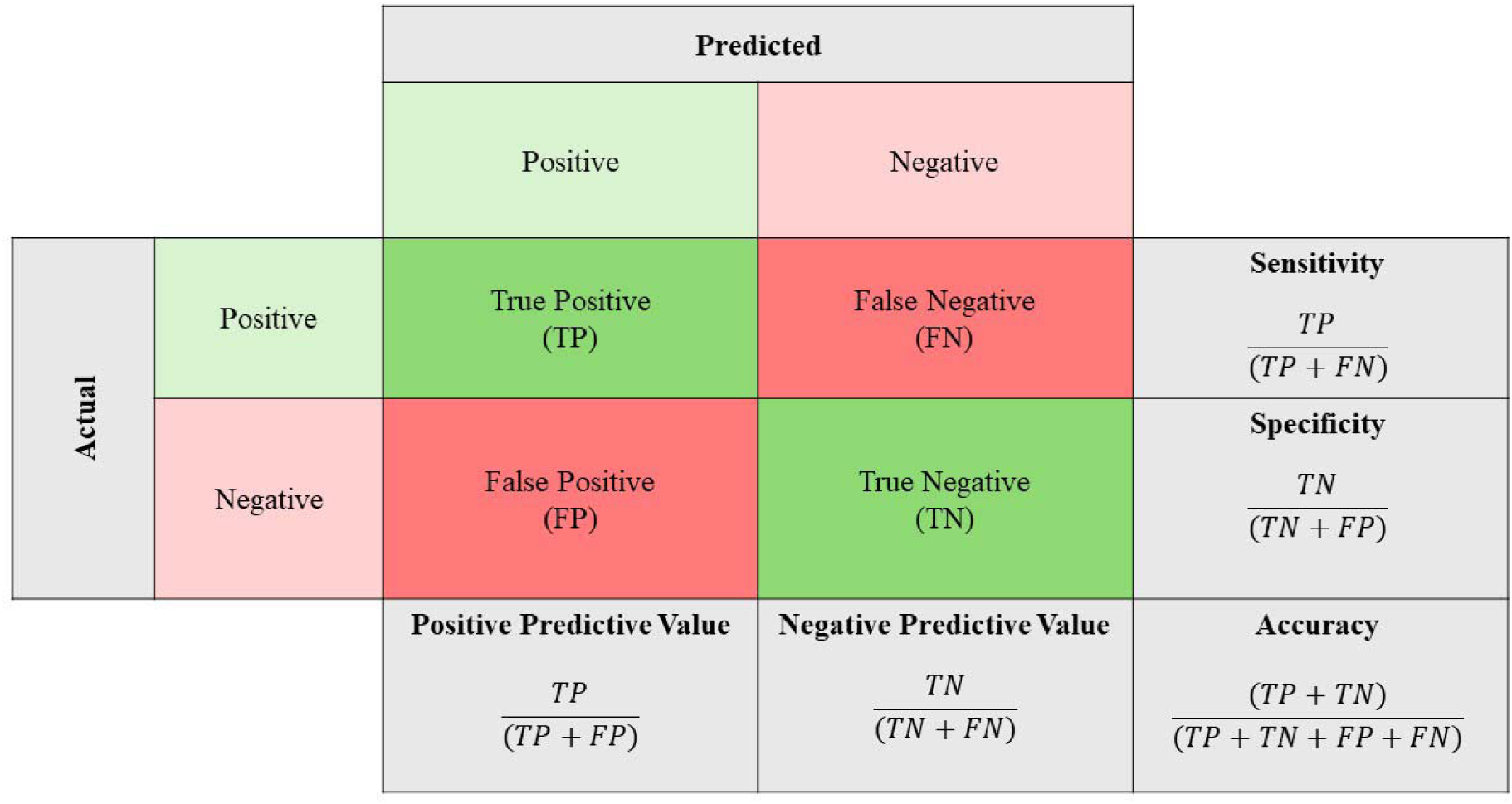
Confusion matrix depicting calculations of sensitivity, specificity, positive predictive value, negative predictive value, and accuracy

**Table 1:** NESARC-III and clinical sample participant characteristics.

| **Demographics**<br>*M(SD)/%/Median* | **NESARC-III**<br>*(N = 25,778)* | **Clinical Sample**<br>*(N=1,341)* |
| --- | --- | --- |
| Sex (% Female) | 53.5 | 32.1 |
| Ethnicity (% White) | 55.5 | 87.1 |
| Age | 43.4 (16.5) | 41.6 (11.7) |
| Education | 10<sup>th</sup> grade | Some college/university |
| **AUD Status (%+)** | 19.9 | 82.4 |
| *Mild*<sup>*</sup> | 51.1 | 8.1 |
| *Moderate*<sup>*</sup> | 23.1 | 11.0 |
| *Severe*<sup>*</sup> | 25.8 | 80.9 |
| **Drinking Characteristics** |  |  |
| Number of Drinking Days<sup>+</sup> | 2 to 3 times per month | --- |
| Drinks per Drinking Day<sup>+</sup> | 3.2 (6.7) | --- |
| Largest Drinks per Drinking Day<sup>+</sup> | 6.0 (10.2) | --- |
| Frequency of 5+ Drinks in One Day<sup>+</sup> | 11 | --- |
| Frequency of 8+ Drinks in One Day<sup>+</sup> | 11 | --- |
| Frequency of 12+ Drinks in One Day<sup>+</sup> | 11 | --- |
| Frequency of 5+ Drinks in ≤ 2 Hours<sup>+</sup> | 11 | --- |
| Frequency of > DLRG<sup>+</sup> | 34 (78.2) | --- |
| Number of Drinking Days<sup>**</sup> | --- | 48.5 (35.2) |
| Drinks per Drinking Day<sup>**</sup> | --- | 10.3 (10.1) |
| Number of Heavy Drinking Days<sup>**</sup> | --- | 41.4 (35.7) |
| Frequency of Alcohol Use<sup>**</sup> | --- | Multiple times weekly |

\* percentage of AUD+ sample, values derived using raw data reported in Grant et al., 2015; <sup>+</sup> = measured over previous year; <sup>\*</sup> = measured over 90 days,

#### Clinical Sample

Participants were adults seeking treatment at a Canadian mental health and addictions inpatient treatment centre in Guelph, Ontario. Consecutive, unique individuals who entered between April 26^th^, 2018, to February 28^th^, 2020, consented to research, and reported any drinking over the past 90 days were included in this study (*N* = 1,341). Among those with AUD, the majority had a severe condition. See Table 1 for participant characteristics and Figure 1 for STARD diagram. Following stabilization (two to seven days following admission), patients completed a self-report measurement-based care battery that included questions about their alcohol consumption in the 90 days prior to entering the treatment facility. All procedures were approved by the Regional Centre for Excellence in Ethics, Research Ethics Board in Guelph, Ontario, Canada (protocol #19-8).

### Measures

#### Community Sample

The reference standard for past year AUD was the Alcohol Use Disorder and Associated Disabilities Interview Schedule Version 5 (AUDADIS-5), a diagnostic interview. Participants were classified as having an AUD if they experienced at least two of the 11 DSM-5 diagnostic criteria at the same time in the past year. Index tests for this sample included the following: (1) <u>Number of drinking days</u>: Participants reported the number of days they consumed at least one alcoholic drink over the previous 12 months, choosing from binned responses: 1 to 2 times in the last year, 3 to 6 times in the last year, 7 to 11 times in the last year, once a month, 2 to 3 times a month, once a week, 2 times a week, 3 to 4 times a week, nearly every day, or every day. (2) <u>Number of drinks per drinking day</u>: Participants reported the average number of drinks they consumed on drinking days over the previous 12 months with a numeric value. (3) <u>Largest number of drinks consumed on a drinking day</u>: Participants reported the largest number of drinks they consumed on a drinking day over the previous 12 months with a numeric value. (4-6) <u>Frequency of drinking over 5, 8, and 12 drinks in one day</u>. Participants reported how often they consumed more than 5/8/12 drinks in a single day over the previous 12 months, choosing from binned responses: every day, nearly every day, 3 to 4 times a week, 2 times a week, once a week, 2 to 3 times a month, once a month, 7 to 11 times in the last year, 3 to 6 times in the last year, 1 to 2 times in the last year, or never in the last year. (7<u>) Frequency of drinking 5+ drinks in 2 hours or less</u>: Participants reported how often they drank 5 or more drinks in 2 hours or less over the previous 12 months, choosing from binned responses: every day, nearly every day, 3 to 4 times a week, 2 times a week, once a week, 2 to 3 times a month, once a month, 7 to 11 times in the last year, 3 to 6 times in the last year, 1 or 2 times in the last year, never in the last year. (8) <u>Frequency of exceeding daily low-risk drinking limits</u>. Participants reported how often over the previous 12 months they exceeded daily low-risk drinking limits (4 + drinks per drinking day for females, 5+ drinks per drinking day for males) with a numeric value between 0 and 365.

#### Clinical Sample

The reference standard for AUD was determined by patient reports on a DSM-5 AUD symptom checklist, where participants dichotomously endorsed or denied the presence of the eleven DSM-5 AUD criteria. This checklist has been validated to closely correspond with a structured clinical interview (20). The index tests included the following: (1) <u>Number of drinking days</u>: Participants reported the number of days they consumed at least one alcoholic drink over the 90 days prior to admission, with response options as a numeric value between 0 and 90. (2) <u>Number of drinks per drinking day:</u> Participants reported a numeric value for the average number of drinks per drinking day, with a maximum of 80 to reflect biological plausibility. (3) <u>Number of heavy drinking days:</u> Participants reported the average number of days they drank more than 4 drinks (for females) and 5 drinks (for males) over the 90 days prior to admission, responding with a numeric value from 0 to 90. Finally, a categorical item was included in the battery to assess drinking frequency. Participants chose from the following responses regarding their drinking frequency over the 90 days prior to admission: “None”, “Once a month”, “Once a week”, “Multiple times weekly”, “Once per day”, or “Multiple times daily”.

### Data Analysis

Receiver operating characteristic (ROC) curves were constructed to examine drinking variables as classifiers of AUD status. ROC curves depict the relationship between the true positive and false positive rate at every possible cut-off value of a given variable (Mandrekar, 2010). Area under the ROC curve (AUC) was used to examine the predictive capability of the drinking variables. An AUC value of 0.5 indicates no discriminatory capability, and a value of 1.0 indicates perfect discriminatory capability (21). For drinking variables that were statistically significant discriminators, the optimal cut-off value was identified via the Youden Index (22).

Subsequently, accuracy, sensitivity, specificity, positive predictive value (PPV), and negative predictive value (NPV) were calculated using this value. Accuracy refers to the percentage of accurate/inaccurate predictions associated with the chosen cut-off. Sensitivity refers to the ability of the test to yield a positive result when AUD is present, while specificity refers to the ability of the test to yield negative results when AUD is absent. PPV refers to the true positive rate among all positive predictions, while NPV refers to the true negative rate among all negative predictions. Both PPV and NPV are impacted by prevalence, with higher prevalence associated with higher PPV and lower NPV, and lower prevalence associated with poorer PPV and higher NPV (23). See Figure 2 for a visualization and mathematical descriptions of these indices. Variables were considered clinically useful if the sum of sensitivity and specificity was greater that 1.5, where 1 represents useless and 2 represents perfect predictions (24).

**Figure 2:**
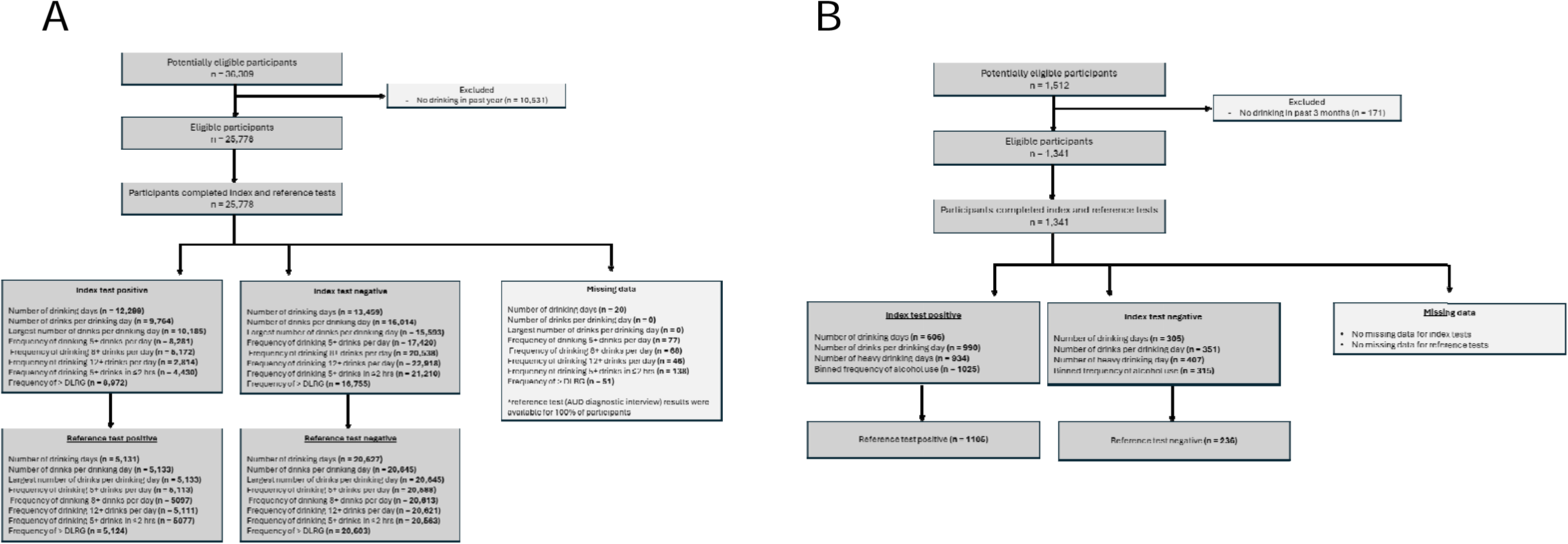
STARD diagram depicting participant flow throw study. Panel A depicts the community adult sample. Panel B depicts the inpatient clinical sample.

These analyses were conducted for each individual drinking variable in both samples. First, analyses were conducted for the overall community sample, followed by the clinical sample. Then, sex specific analyses were conducted for both samples to examine potential differences by sex. All analyses had sufficient power to detect significant cut-offs (25). There was no missing data for the reference standard (i.e., AUD status) in either sample. Participants missing data for an index text (i.e., drinking quantity/frequency variable) were excluded from that particular analysis. See Supplemental Materials for a cross tabulation of the index text results by results from the reference standard. ROC analyses were carried out using SPSS version 26 (26). Accuracy indices (e.g., sensitivity, specificity, PPV, NPV, accuracy, and error rates) were calculated in R, version 4.1.2 (27) using the pROC package (28).

## RESULTS

### Overall Analyses

#### Community Sample

Table 2 presents the AUC, their associated confidence intervals, optimal cut-offs, and corresponding accuracy indices of each drinking variable. Results in the overall sample revealed that all the drinking variables performed significantly better than chance at classifying AUD status (AUCs = .66-.86; *p*s<.0001). Overall sample accuracy ranged from 0.65 for Number of drinking days to 0.84 for Frequency of drinking 12+ drinks in one day. Across most variables, sensitivity and specificity were generally balanced, ranging 0.72 to 0.84. However, sensitivity and specificity were unbalanced for Number of drinking days, Frequency of drinking 8+/12+ drinks in a single day, and Frequency of drinking 5+ drinks in 2 hours or less, with low sensitivity (0.37-0.61) and high specificity (0.81-0.95). PPV was low across variables, ranging 0.34 (number of drinking days) to 0.66 (frequency of 12+ drinks in one day). NPV was excellent, ranging 0.86 to 0.95 (frequency of 12+ drinks in one day, largest drinks per drinking day, respectively). Only three variables surpassed the clinical utility threshold: Largest drinks per drinking day, Frequency of drinking 5+ drinks in one day, and Frequency of exceeding daily low risk drinking guidelines (1.56-1.58).

**Table 2:** NESARC and clinical sample AUC, optimal cut-offs, and accuracy indices in overall sample and by sex.

| Measure | Optimal cut-off | AUC, SE (95% CI) | Accuracy | Sens. | Spec. | PPV | NPV | Sens. + Spec. |
| --- | --- | --- | --- | --- | --- | --- | --- | --- |
| **Community Sample** |  |  |  |  |  |  |  |  |
| Number of drinking days | ≥2-3x per month | **.77**, .003 (.77-.78) | .65 | .81 | .61 | .34 | .93 | 1.42 |
| Female | ≥2-3 per month | **.79**, .005 (.78-.80) | .69 | .76 | .68 | .31 | .94 | 1.44 |
| Male | ≥1 per week | **.74**, .005 (.74-.75) | .67 | .73 | .64 | .40 | .88 | 1.37 |
| Number of drinks per drinking day | ≥2 | **.80**, .003 (.80-.81) | .73 | .77 | .72 | .40 | .93 | 1.49 |
| Female | ≥2 | **.80**, .003 (.80-.81) | .78 | .70 | .79 | .39 | .93 | 1.49 |
| Male | ≥2 | **.79**, .005 (.78-.79) | .67 | .82 | .62 | .42 | .91 | 1.44 |
| Largest drinks per drinking day | ≥4 | **.86**, .003 (.85-.86) | .74 | .84 | .72 | .42 | .95 | **1.56** |
| Female | ≥3 | **.87**, .004 (.86-.87) | .72 | .88 | .69 | .35 | .97 | **1.57** |
| Male | ≥5 | **.84**, .004 (.83-.85) | .72 | .85 | .68 | .46 | .93 | **1.53** |
| Frequency of 5+ drinks in one day | ≥1-2x per year | **.83**, .003 (.83-.84) | .79 | .77 | .79 | .48 | .93 | **1.56** |
| Female | ≥never | **.82**, .006 (.80-.83) | .80 | .76 | .81 | .43 | .95 | **1.57** |
| Male | ≥3-6x per year | **.83**, .004 (.83-.84) | .77 | .79 | .76 | .52 | .92 | **1.55** |
| Frequency of 8+ drinks in one day | ≥never | **.75**, .004 (.74-.76) | .83 | .59 | .89 | .58 | .90 | 1.48 |
| Female | ≥never | **.70**, .007 (.69-.71) | .87 | .44 | .95 | .63 | .90 | 1.39 |
| Male | ≥never | **.78**, .006 (.77-.79) | .79 | .69 | .82 | .56 | .89 | **1.51** |
| Frequency of 12+ drinks in one day | ≥never | **.66**, .005 (.65-.67) | .84 | .37 | .95 | .66 | .86 | 1.32 |
| Female | ≥never | **.60**, .007 (.58-.61) | .86 | .21 | .98 | .71 | .87 | 1.19 |
| Male | ≥never | **.70**, .006 (.69-.71) | .81 | .48 | .91 | .65 | .84 | 1.39 |
| Frequency of 5+ drinks in < 2 hours | ≥never | **.71**, .005 (.70-.72) | .83 | .50 | .91 | .57 | .88 | 1.41 |
| Female | ≥never | **.68**, .007 (.66-.69) | .86 | .40 | .95 | .59 | .89 | 1.35 |
| Male | ≥never | **.73**, .006 (.72-.74) | .79 | .58 | .86 | .57 | .86 | 1.44 |
| Frequency of > DLRG | ≥6x per year | **.85**, .003 (.85-.86) | .77 | .81 | .77 | .46 | .94 | **1.58** |
| Female | ≥3x per year | **.85**, .005 (.85-.86) | .74 | .86 | .72 | .37 | .97 | **1.58** |
| Male | ≥6x per year | **.84**, .004 (.84-.85) | .75 | .83 | .73 | .50 | .93 | **1.56** |
| **Clinical Sample** |  |  |  |  |  |  |  |  |
| Drinking days | ≥16 | **.86**, .012 (.84-.89) | .78 | .77 | .83 | .95 | .44 | **1.60** |
| Female | ≥15 | **.86**, .025 (.82-.91) | .77 | .76 | .88 | .98 | .35 | **1.64** |
| Male | ≥17 | **.87**, .014 (.84-.92) | .79 | .78 | .81 | .94 | .48 | **1.59** |
| Drinks per drinking day | ≥3 | **.89**, .014 (.86-.91) | .85 | .86 | .82 | .96 | .55 | **1.68** |
| Female | ≥4 | **.85**, .034 (.79-.92) | .82 | .83 | .80 | .97 | .41 | **1.63** |
| Male | ≥4 | **.90**, .015 (.87-.93) | .87 | .87 | .83 | .95 | .62 | **1.70** |
| Heavy drinking days | ≥6 | **.92**, .010 (.90-.94) | .85 | .84 | .86 | .97 | .54 | **1.71** |
| Female | ≥3 | **.92**, .022 (.87-.96) | .89 | .90 | .80 | .97 | .56 | **1.70** |
| Male | ≥5 | **.92**, .011 (.90-.94) | .85 | .85 | .87 | .96 | .59 | **1.72** |
| Binned frequency of alcohol use | ≥once a week | **.89**, .012 (.87-.92) | .88 | .89 | .81 | .96 | .61 | **1.70** |
| Female | ≥once a week | **.91**, .024 (.86-.95) | .88 | .87 | .89 | .98 | .52 | **1.76** |
| Male | ≥once a week | **.89**, .014 (.86-.92) | .87 | .90 | .79 | .95 | .65 | **1.69** |

AUC = Area under the curve, DLRG = Daily Low Risk Drinking Guidelines, Sens + Spec = sensitivity + specificity, PPV = Positive Predictive Value, NPV = Negative Predictive Value. Bolded AUC values = $p < .0001$ ; Bolded Sens + Spec values indicate clinically useful indicators.

#### Clinical Sample

Table 2 presents the AUC, associated confidence intervals, optimal cut-offs, and corresponding accuracy indices of each drinking variable. All drinking variables performed significantly better than chance at classifying AUD status (AUCs = .86-.92; *p*s<.001). A cut-off of 16+ drinking days correctly classified AUD status in 78% of cases and was associated with moderate to high sensitivity and specificity (0.77 and 0.83, respectively), high positive predictive power, but low negative predictive power (0.94 and 0.44, respectively). A cut-off of drinking 3+ drinks per drinking day correctly classified AUD status in 85% of cases. This cut-off was associated with high sensitivity and specificity (0.86 and 0.82, respectively), high positive predictive power, but low levels of negative predictive power (0.96 and 0.55, respectively). A cut-off of 6+ heavy drinking days correctly classified AUD status in 83% of cases and was associated with high sensitivity and specificity (0.82 and 0.89, respectively), high positive predictive power, and low negative predictive power (0.97 and 0.51, respectively). A cut-off of drinking 1x per week or more correctly classified AUD status in 88% of cases. This cut-off was associated with high sensitivity and specificity (0.89 and 0.81, respectively), high positive predictive power but low negative predictive power (0.96 and 0.61, respectively). All variables surpassed the clinical utility threshold (1.60-1.71).

### Sex Differences

#### Community Sample

All drinking variables performed significantly better than chance at classifying AUD status across both males (AUCs = 0.70-0.84, *p*s<.0001) and females (AUCs = 0.60-0.87, *p*s<.0001). In the community sample, cutoffs differed among males and females for (1) Number of drinking days (2-3x per months or more for females, once per week or more for males), (2) Largest drinks per drinking day (3 for females, 5 for males), (3) Frequency of drinking 5+ drinks per day (once or more for females, 6 or more times for males), and (4) Frequency of exceeding daily low risk guidelines (3 or more for females, 6 or more for males). Regarding clinical utility, the same heavy drinking metrics emerged as clinically useful indicators as the overall sample. However, Frequency of drinking 8+ drinks per day surpassed the clinical utility threshold for males (1.51), but not females.

#### Clinical Sample

All drinking variables performed significantly better than chance at classifying AUD status across both males (AUCs = 0.87-0.92, *p*s<.0001) and females (AUCs = 0.85-0.92, *p*s<.0001). Again, optimal cut-offs were largely similar across sexes, with differences for (1) Drinking days (15 or more for females, 17 or more for males), and (2) Heavy drinking days (3 or more for females, 5 or more for males). Across accuracy indices, values were generally similar across sexes and all indices remained clinically useful for both sexes.

## DISCUSSION

This study used ROC curves and a variety of classification indices to investigate whether alcohol consumption characteristics validly identified AUD status among two samples, a large nationally representative sample of community adults in the United States and a sample of patients seeking inpatient substance use disorder treatment in Canada. Across samples and sexes, analyses revealed that drinking characteristics performed much better than chance at classifying AUD status. In examining sensitivity and specificity values, several indicators surpassed the 1.5 threshold of clinical utility.

Specifically, among both samples, measures of heavy drinking emerged as clinically useful indicators. In the community sample, only Largest number of drinks per drinking day, Frequency of drinking 5+ drinks per day, and Frequency of exceeding daily low risk guidelines emerged as clinically useful indicators in the overall sample. In the overall clinical sample, all drinking indicators passed the thresholds for clinical utility. However, the measure of heavy drinking (i.e., 6+ heavy drinking days over the previous 90 days) emerged as a particularly discriminating variable. Together, findings point towards the utility of heavy drinking as a clinically useful metric in helping confer a diagnosis of AUD.

Sex specific analyses revealed that sex did not substantially impact the classificatory capacity of drinking behaviour at classifying AUD status. Optimal cut-offs were largely similar across sexes, with slight differences in few variables. When differences did occur, cut-offs were slightly elevated for males compared to females. Further, accuracy indicie values were generally similar across males and females. Regarding clinically informative metrics, those that were clinically informative in the overall sample remained so for both sexes. In addition, Frequency of drinking 8+ drinks in one day surpassed the 1.50 threshold (1.51) for males. Taken together, findings suggest that despite several small differences in cut-offs, drinking metrics generally operate with a similar degree of accuracy across both males and females.

Results from this study are consistent with studies that found the AUDIT-C (a compilation of three quantity/frequency items scored on a Likert scale) performed well as a screener for AUD status. However, this study extends these findings and suggests that even a single item of frequency of heavy drinking can classify AUD status with considerable accuracy, without needing to score and interpret a scale. Indeed, individual items may be a particularly useful tool in clinical settings, given the ease of administration, heuristic value, and lack of scoring required. This study suggests that heavy drinking frequency is a clinically useful tool for diagnosing AUD. These variables may hold particular value in a measurement-based care system (29), where alcohol consumption is routinely collected and evaluated as a marker of treatment progress, and as a possible tool to complement existing diagnostic definitions. Importantly, however, we are not proposing that quantity/frequency measures be used in isolation in diagnostic decisions. Rather, measures of heavy drinking may serve as an effective, easy-to-administer tool for clinicians to use in conjunction with other validated instruments. Of note, this study’s findings empirically add to the discourse about whether AUD can be fundamentally thought of as “heavy drinking over time” (3,4,6). Here, the measures of heavy drinking frequency were the variables that most robustly and precisely classified AUD status, suggesting that heavy use over time may not be synonymous with AUD, but can serve as a useful clinical indicator for AUD status.

These findings must be considered within the context of the study’s strengths and limitations. Both samples used to conduct analyses were large, with sufficient power to detect effects (25). Findings from analyses conducted with the community adult population are generalizable to the general community of drinking adults, due to the representative nature of the sample. Additionally, the reference standard in both samples were of high quality: a diagnostic interview in the community adult sample and patient self-report of the eleven DSM-5 criteria in the clinical sample. A limitation was that AUD status was not based on a clinical interview in the clinical sample, but this is mitigated by evidence that self-report closely corresponds with responses to clinical interviews in clinical samples where there are no contingencies favoring minimization (20). Another limitation is that NESARC-III did not collect participant gender identity, and as such this study was unable to examine possible differences by gender identity in addition to sex. Thus, it is possible that findings from this study may not generalize to gender diverse individuals. Nonetheless, sex analyses in this study support that alcohol consumption characteristics accurately classify AUD among both males and females.

To conclude, this study’s findings broadly suggest that patterns of alcohol consumption can classify AUD status with considerable accuracy. Specifically, measures of heavy drinking frequency emerged as robust indicators, surpassing benchmarks for utility in clinical practice. Analyses from this study suggest that these findings are relevant across both male and female community adult samples, as well as in a treatment seeking clinical sample. This work broadly supports the potential utility of quantitative indicators of alcohol consumption in conferring a diagnosis of AUD.

## Data Availability

Data produced in the present study may be made available upon reasonable request to the authors

## Notes

**Funding Acknowledgments:** Funding to support this work was provided by the Peter Boris Chair in Addictions Research, and a Canada Research Chair in Translational Addiction Research (CRC-2020-00170)

### Competing Interest Statement

James MacKillop is a principal in BEAM Diagnostics, Inc. and has served as a consultant to Clairvoyant Therapeutics. No other authors have disclosures.

### Funding Statement

Funding to support this work was provided by the Peter Boris Chair in Addictions Research, a Canada Research Chair in Translational Addiction Research (CRC-2020-00170), and Homewood Research Institute, a registered charity.

### Author Declarations

The Regional Centre for Excellence in Ethics, Research Ethics Board in Guelph, Ontario, Canada gave ethical approval for this work (protocol #19-8).

