## supplemental for "Diagnostic Validity of Drinking Behaviour for Identifying Alcohol Use Disorder: Findings from a Nationally Representative Sample of Community Adults and an Inpatient Clinical Sample": Supplemental Materials.pdf

### Clinical Sample Crosstabulations

Drinking Days

Full Sample

| | | Predicted | |
|---|---|---|---|
| | | Positive | Negative |
| Actual | Positive | 855 | 250 |
| | Negative | 41 | 195 |

Female Sample

| | | Predicted | |
|---|---|---|---|
| | | Positive | Negative |
| Actual | Positive | 283.87 | 90.13 |
| | Negative | 7 | 49 |

Male Sample

| | | Predicted | |
|---|---|---|---|
| | | Positive | Negative |
| Actual | Positive | 571.64 | 159.36 |
| | Negative | 34.02 | 145.98 |

Drinks per Drinking Day

Full Sample

| | | Predicted | |
|---|---|---|---|
| | | Positive | Negative |
| Actual | Positive | 948.09 | 156.91 |
| | Negative | 42.01 | 193.99 |

Female Sample

| | | Predicted | |
|---|---|---|---|
| | | Positive | Negative |
| Actual | Positive | 309 | 65 |
| | Negative | 11 | 45 |

Male Sample

| | | Predicted | |
|---|---|---|---|
| | | Positive | Negative |
| Actual | Positive | 639 | 92 |
| | Negative | 31 | 149 |

Heavy Drinking Days

Full Sample

| | | Predicted | |
|---|---|---|---|
| | | Positive | Negative |
| Actual | Positive | 930 | 175 |
| | Negative | 32 | 204 |

Female Sample

| | | Predicted | |
|---|---|---|---|
| | | Positive | Negative |
| Actual | Positive | 338.10 | 35.90 |
| | Negative | 10.98 | 45.02 |

Male Sample

| | | Predicted | |
|---|---|---|---|
| | | Positive | Negative |
| Actual | Positive | 621.35 | 109.65 |
| | Negative | 23.04 | 156.96 |

Binned Alcohol Use Frequency

Full Sample

| | | Predicted | |
|---|---|---|---|
| | | Positive | Negative |
| Actual | Positive | 981.24 | 123.76 |
| | Negative | 43.90 | 192.10 |

Female Sample

| | | Predicted | |
|---|---|---|---|
| | | Positive | Negative |
| Actual | Positive | 326.88 | 47.12 |
| | Negative | 5.99 | 50.01 |

Male Sample

| | | Predicted | |
|---|---|---|---|
| | | Positive | Negative |
| Actual | Positive | 654.25 | 76.76 |
| | Negative | 37.98 | 142.02 |

### Community Sample Crosstabulations

Number of drinking days

Full Sample

| | | Predicted | |
|---|---|---|---|
| | | Positive | Negative |
| Actual | Positive | 4,150.98 | 980.02 |
| | Negative | 8,147.67 | 12,479.34 |

N missing = 20

Female Sample

| | | Predicted | |
|---|---|---|---|
| | | Positive | Negative |
| Actual | Positive | 1,654.41 | 519.59 |
| | Negative | 3,768.05 | 7,825.95 |

N missing = 11

Male Sample

| | | Predicted | |
|---|---|---|---|
| | | Positive | Negative |
| Actual | Positive | 2167.48 | 3215.75 |
| | Negative | 789.52 | 5817.25 |

N missing = 9

Number of drinks per drinking day

Full Sample

| | | Predicted | |
|---|---|---|---|
| | | Positive | Negative |
| Actual | Positive | 3,942.14 | 1,190.86 |
| | Negative | 5,821.89 | 14,823.11 |

Female Sample

| | | Predicted | |
|---|---|---|---|
| | | Positive | Negative |
| Actual | Positive | 1,515.98 | 659.03 |
| | Negative | 2,425.24 | 9,178.76 |

Male Sample

| | | Predicted | |
|---|---|---|---|
| | | Positive | Negative |
| Actual | Positive | 2,425.56 | 3,408.46 |
| | Negative | 532.44 | 5,632.54 |

Largest number of drinks per drinking day

Full Sample

| | | Predicted | |
|---|---|---|---|
| | | Positive | Negative |
| Actual | Positive | 4,321.99 | 811.01 |
| | Negative | 5,863.18 | 14,781.82 |

Female Sample

| | | Predicted | |
|---|---|---|---|
| | | Positive | Negative |
| Actual | Positive | 1,911.83 | 263.18 |
| | Negative | 3,550.82 | 8,053.18 |

Male Sample

| | | Predicted | |
|---|---|---|---|
| | | Positive | Negative |
| Actual | Positive | 2,505.43 | 2,902.16 |
| | Negative | 452.57 | 6,138.84 |

Frequency of 5+ drinks in one day

Full Sample

| | | Predicted | |
|---|---|---|---|
| | | Positive | Negative |
| Actual | Positive | 3,957.46 | 1,155.54 |
| | Negative | 4,323.48 | 16,264.52 |

N missing = 77

Female Sample

| | | Predicted | |
|---|---|---|---|
| | | Positive | Negative |
| Actual | Positive | 1,648.97 | 515.03 |
| | Negative | 2,200.39 | 9,380.61 |

N missing = 34

Male Sample

| | | Predicted | |
|---|---|---|---|
| | | Positive | Negative |
| Actual | Positive | 2,323.81 | 625.19 |
| | Negative | 2,152.67 | 6,854.33 |

N missing = 43

Frequency of 8+ drinks in one day

Full Sample

| | | Predicted | |
|---|---|---|---|
| | | Positive | Negative |
| Actual | Positive | 2,986.84 | 2,110.16 |
| | Negative | 2,184.98 | 18,428.02 |

N missing = 68

Female Sample

| | | Predicted | |
|---|---|---|---|
| | | Positive | Negative |
| Actual | Positive | 947.36 | 1,210.64 |
| | Negative | 568.06 | 11,024.94 |

N missing = 28

Male Sample

| | | Predicted | |
|---|---|---|---|
| | | Positive | Negative |
| Actual | Positive | 2,039.67 | 899.33 |
| | Negative | 1,623.60 | 7,396.40 |

N missing = 40

Frequency of 12+ drinks in one day

Full Sample

| | | Predicted | |
|---|---|---|---|
| | | Positive | Negative |
| Actual | Positive | 1,865.52 | 3,245.49 |
| | Negative | 948.57 | 19,672.43 |

N missing = 46

Female Sample

| | | Predicted | |
|---|---|---|---|
| | | Positive | Negative |
| Actual | Positive | 454.86 | 1,711.14 |
| | Negative | 185.55 | 11,411.45 |

N missing = 16

Male Sample

| | | Predicted | |
|---|---|---|---|
| | | Positive | Negative |
| Actual | Positive | 1,407.71 | 1,537.29 |
| | Negative | 776.06 | 8,247.94 |

N missing = 30

Frequency of 5+ drinks in<2 hours

Full Sample

| | | Predicted | |
|---|---|---|---|
| | | Positive | Negative |
| Actual | Positive | 2,538.50 | 2,538.50 |
| | Negative | 1,891.80 | 18,671.20 |

N missing = 138

Female Sample

| | | Predicted | |
|---|---|---|---|
| | | Positive | Negative |
| Actual | Positive | 849.65 | 1,301.36 |
| | Negative | 601.54 | 10,966.46 |

N missing = 60

Male Sample

| | | Predicted | |
|---|---|---|---|
| | | Positive | Negative |
| Actual | Positive | 1,688.30 | 1,237.70 |
| | Negative | 1,295.28 | 7,699.72 |

N missing = 78

Frequency of> DLRG

Full Sample

| | | Predicted | |
|---|---|---|---|
| | | Positive | Negative |
| Actual | Positive | 4,150.44 | 973.56 |
| | Negative | 4,821.10 | 15,781.90 |

N missing = 51

Female Sample

| | | Predicted | |
|---|---|---|---|
| | | Positive | Negative |
| Actual | Positive | 1,867.51 | 301.49 |
| | Negative | 3,220.91 | 8,365.09 |

N missing = 24

Male Sample

| | | Predicted | |
|---|---|---|---|
| | | Positive | Negative |
| Actual | Positive | 2,452.65 | 502.35 |
| | Negative | 2,461.64 | 6,555.36 |

N missing = 27
